# Prevalence and Mortality Trends of Hypertension Subtypes Among US Adults: An Analysis of the National Health and Nutrition Examination Survey (NHANES) 1999-2018

**DOI:** 10.1101/2024.09.27.24314521

**Authors:** Kevin S Tang, Jeffrey E Jones, Wenjun Fan, Nathan D Wong

**Affiliations:** Heart Disease Prevention Program, Mary and Steve Wen Cardiovascular Division, Department of Medicine, University of California, Irvine, CA; Department of Epidemiology and Biostatistics, University of California, Irvine, CA

**Author notes:** Authors’ Contributions: KST and JEJ contributed to writing, editing, and data analysis. NDW conceived of the project. NDW and WF contributed to editing and oversight. Each author contributed important intellectual content during manuscript drafting or revision and accepts accountability for the overall work by ensuring that questions pertaining to the accuracy or integrity of any portion of the work are appropriately investigated and resolved. Corresponding Author: Nathan D. Wong, PhD, Heart Disease Prevention Program, C240 Medical Sciences, University of California Irvine, Irvine, CA 92697.

**Keywords:** Hypertension, Blood Pressure, Cardiovascular Disease

## Abstract

Hypertension (HTN) has been demonstrated as one of the leading risk factors for development of cardiovascular disease (CVD) and CVD mortality. This study examines the prevalence and distribution of HTN subtypes (isolated diastolic hypertension [IDH], isolated systolic hypertension [ISH], and systolic-diastolic hypertension [SDH]) across age, sex, and race/ethnicity per the nationally representative National Health and Nutrition Examination Survey (NHANES) from 1999 to 2018 based on the updated 2017 ACC/AHA HTN definition and their association with CVD and all-cause mortality. Among US adults, the overall prevalence of HTN is 41.1%. Across increasing age, the prevalence of IDH decreased, ISH increased, and SDH increased and peaked in the 6^th^ decade of life after which SDH prevalence decreased. By age 80, over 80% of persons with HTN demonstrated ISH. A sub-cohort from NHANES 1999-2008 with follow-up until 2018 showed that ISH and SDH were most strongly associated with increased risk for CVD (HR 1.20 and 1.37, p<0.01, respectively) and all-cause mortality (HR 1.17 and 1.21, p<0.01, respectively). Our data demonstrate the continuing importance of HTN subtype transitions across age and their differences in predicting future CVD and total mortality.

---

Hypertension (HTN) has long been demonstrated to be an independent predictor of cardiovascular disease (CVD) and all-cause mortality.^1–3^ Elevated systolic blood pressure (SBP) has been particularly associated with worse CVD outcomes when compared to elevated diastolic blood pressure (DBP) alone.^2,4,5^ Thus, the presence of specific subtypes of HTN may have variable implications for risk of mortality. The Ohasama Study provided high-quality data demonstrating that systolic HTN conferred significantly increased risk of CVD mortality while isolated diastolic hypertension did not portend a higher risk when compared to normotension.^6^ Further analyses have suggested that cardiovascular death risk modulation by hypertension subtypes are unequally affected by age and sex, and that different subtypes may confer variable risks in different subject cohorts.^7^ In 2017, the Global Burden of Disease Study found high SBP to be the leading factor for attributable deaths and disability-adjusted life-years (DALYs), accounting for 10.4 million deaths and 218 million DALYs globally in the year of 2017 alone.^8^ This correlation is increasingly salient as the prevalence of HTN in the US adult population has steadily increased from around 24% in 2001^2^ to over 30% in 2018 (using the pre-2017 cutoff of 140/90 mmHg).^9^ A separate analysis of the Global Burden of Disease Study found that between 1990 and 2019, the number of deaths directly attributable to high SBP increased by 54.1% with a concurrent increase in number of DALYs of 52.4%.^10^ However, despite the strength of evidence for the cardiovascular benefits of strict HTN treatment^11^, data suggests that less than 40% of hypertensive adults in the US are treated to blood pressure goal.^2,12^

In the early 2000s, Franklin et al. characterized the prevalence of HTN and its subtypes in the US Adult Population using the National Health and Nutrition Examination Survey (NHANES).^2^ The purpose of this study is to re-evaluate the prevalence of HTN subtypes according to the more recent blood pressure cut points based on the 2017 American College of Cardiology (ACC)/American Heart Association (AHA) Guidelines for High Blood Pressure^13^ stratifying by sex and race/ethnicity. We also examined these HTN subtypes in relation to other comorbidities, CVD, and total mortality, respectively, among US adults.

## Methods

### Data Source

This study utilized publicly available de-identified data and was thus exempt from institutional review board review. The NHANES is a cross-sectional survey administered by the Centers for Disease Control and Prevention with the stated goal of collecting demographic information, medical history, and medical examination data on the noninstitutionalized civilian US population. Individual-level demographic characteristics, health and nutrition information, and physical exam and laboratory data were collected through personal interviews and a standardized physical examination conducted in a mobile examination center. Since 1999, sample methodology has consisted of multiyear, stratified, clustered four-stage samples, targeting around 5000 individuals in 15 counties during 2-year periods.^14^ Our analysis included US adults aged 20 or older from NHANES years 1999 – 2018 with valid blood pressure measurements. For mortality analyses we included a baseline cohort from 1999-2008 with CVD and all-cause mortality follow-up through 2018.

### Definitions

Per the 2017 ACC/AHA Blood Pressure Guidelines, HTN was defined as SBP ≥130 mmHg or DBP ≥80 mmHg or those currently on antihypertensive therapy.^13,15^ A SBP 130-139 mmHg or DBP 80-89 mmHg constituted Stage I HTN and SBP ≥140 mmHg or DBP ≥90 mmHg was considered Stage II HTN. Elevated blood pressure was further defined as those with SBP 120-129 mmHg and DBP <80 mmHg. Treatment of HTN was defined as patient-reported use of a prescription antihypertensive agent at the time of the interview.

Subject data were stratified by age (<40, 40-49, 50-59, 60-69, 70-79, and ≥80 years old, respectively), sex, ethnicity (categorized into Hispanic, non-Hispanic White, non-Hispanic Black, and Other including Multi-Ethnic), tobacco use (categorized into current, former, and never smokers), hypertensive subtype, and presence of certain medical comorbidities including obesity, dyslipidemia, diabetes mellitus (DM), and chronic kidney disease (CKD). Dyslipidemia was defined by total cholesterol ≥200 mg/dL, low-density lipoprotein ≥130 mg/dL, high-density lipoprotein <35 mg/dL, or triglycerides ≥150 mg/dL. DM was defined as fasting glucose >126 mg/dL, non-fasting glucose ≥200 mg/dL, hemoglobin A1C ≥6.5%, currently taking insulin or diabetic medication, or those that have been told by a physician that they have DM. CKD was determined through application of the CKD-EPI equation to calculate estimated glomerular filtration rate (eGFR) using serum creatinine, sex, and age. Presence of CKD was defined as eGFR <60 mL/min/1.73m^2^. Three hypertensive subtypes were considered as part of this analysis: isolated systolic hypertension (ISH) defined as SBP ≥130 mmHg and DBP <80 mmHg, isolated diastolic hypertension (IDH) defined as DBP ≥80 mmHg and SBP <130 mmHg, and systolic-diastolic hypertension (SDH) defined as SBP ≥130 mmHg and DBP ≥80 mmHg. Obesity was defined in relation to body mass index (BMI), with BMI 18.5-24.9 kg/m^2^ considered normal weight, BMI 25-29.9 kg/m^2^ considered overweight, and BMI ≥30 kg/m^2^ considered obese. Abdominal obesity, defined as waist circumference >102 cm in men or >88 cm in women, was also used as an indicator of visceral adiposity and examined independently for association with prevalence of hypertensive subtypes. Presence of DM was based on self-report, glucose ≥126 mg/dL if fasting or ≥200 mg/dL if non-fasting, use of DM medications or insulin, or HbA1c≥6.5%. Finally, CKD was defined as estimated glomerular filtration rate <60 mL/min calculated using the 2021 CKD-EPI Creatinine Formula.^16^

### Statistical Analysis

NHANES data were weighted and extrapolated to estimate the prevalence of hypertensive subtypes in the adult non-institutionalized US population. The incidence of cardiovascular disease (CVD) and all-cause mortality was examined and compared between hypertension subtypes against patients with normal blood pressure using Cox regression analysis, adjusted for age, ethnicity, sex, tobacco use, use of antihypertensive agents, and comorbidities including diabetes mellitus, chronic kidney disease, and dyslipidemia.

Mortality data from the National Death Index was linked to NHANES datasets and classified according to ICD-10 codes. CVD death was defined as ICD-10 codes of I00-I09, I11, I13, I20-I51, and I60-I69. Statistical analyses were performed with SAS statistical software (version 9.4; SAS Institute, Cary, NC).

## Results

### Burden of Hypertension and its Subtypes

A total of 72,951 individuals were surveyed in NHANES years 1999–2018, out of which 49,723 had valid blood pressure readings and were included in the analysis. Of these, 41.1% (20,453 individuals projected to 97 million US adults) met criteria for HTN per ACC/AHA guidelines. Of hypertensive adults, 49% had ISH, 32% had SDH, and 19% had IDH (Table 1). Individuals aged 50 or older comprised 60% of all hypertensive adults. Compared to hypertensive adults under the age of 50, individuals aged 50 years or older with hypertension maintained a significantly higher prevalence of ISH (62% versus 22%, P<0.0001). Diastolic HTN, whether as IDH or together with systolic HTN in SDH, comprised the majority of HTN subtypes in adults up to age 59, constituting 74%, 81%, and 65% of HTN in the <40, 40-49, and 50-59 age groups, respectively. Meanwhile, ISH rapidly became the predominant HTN subtype above the age of 60, comprising 85% of all HTN in adults aged 80+. In adults aged 50 or older with hypertension, average SBP was 146 mmHg compared to 132 mmHg in hypertensive adults younger than 50 (p<0.0001) while average DBP values were 74 mmHg and 83 mmHg, respectively (p<0.0001). Figure 1 showed the prevalence of HTN subtypes across age groups.

**Table 1.** Prevalence of Hypertension Subtypes and Demographic Characteristics Among US Adults with Hypertension, NHANES 1999-2018.

|  | **Isolated Systolic Hypertension**<br>**(n=10,083)** | **Isolated Diastolic Hypertension**<br>**(n=3,931)** | **Systolic-Diastolic Hypertension**<br>**(n=6,439)** |
| --- | --- | --- | --- |
| **Total (N=20,453)** | **49.3%** | **19.2%** | **31.5%** |
| **Age** |  |  |  |
| <40 | 4.2% | 7.3% | 4.7% |
| 40-49 | 3.0% | 6.1% | 6.8% |
| 50-59 | 6.2% | 3.7% | 7.8% |
| 60-69 | 13.6% | 1.7% | 7.5% |
| 70-79 | 12.7% | 0.3% | 3.1% |
| 80+ | 9.6% | 0.1% | 1.6% |
| **Sex** |  |  |  |
| Female | 25.5% | 7.3% | 13.8% |
| **Ethnicity** |  |  |  |
| Hispanic | 11.8% | 4.2% | 7.0% |
| White | 23.3% | 8.7% | 11.9% |
| Black | 11.0% | 4.1% | 9.7% |
| Other | 3.2% | 2.3% | 2.9% |
| **Tobacco Use** |  |  |  |
| Current | 7.8% | 4.2% | 7.0% |
| Former | 16.3% | 4.2% | 8.4% |
| **Body Mass Index** |  |  |  |
| <30 | 19.5% | 8.3% | 14.6% |
| 30+ | 29.6% | 10.9% | 17.0% |
| **Dyslipidemia** | 46.5% | 17.8% | 29.8% |
| **Diabetes Mellitus** | 14.6% | 2.1% | 5.9% |
| **Chronic Kidney Disease** | 11.6% | 1.4% | 4.6% |

**Figure 1.**
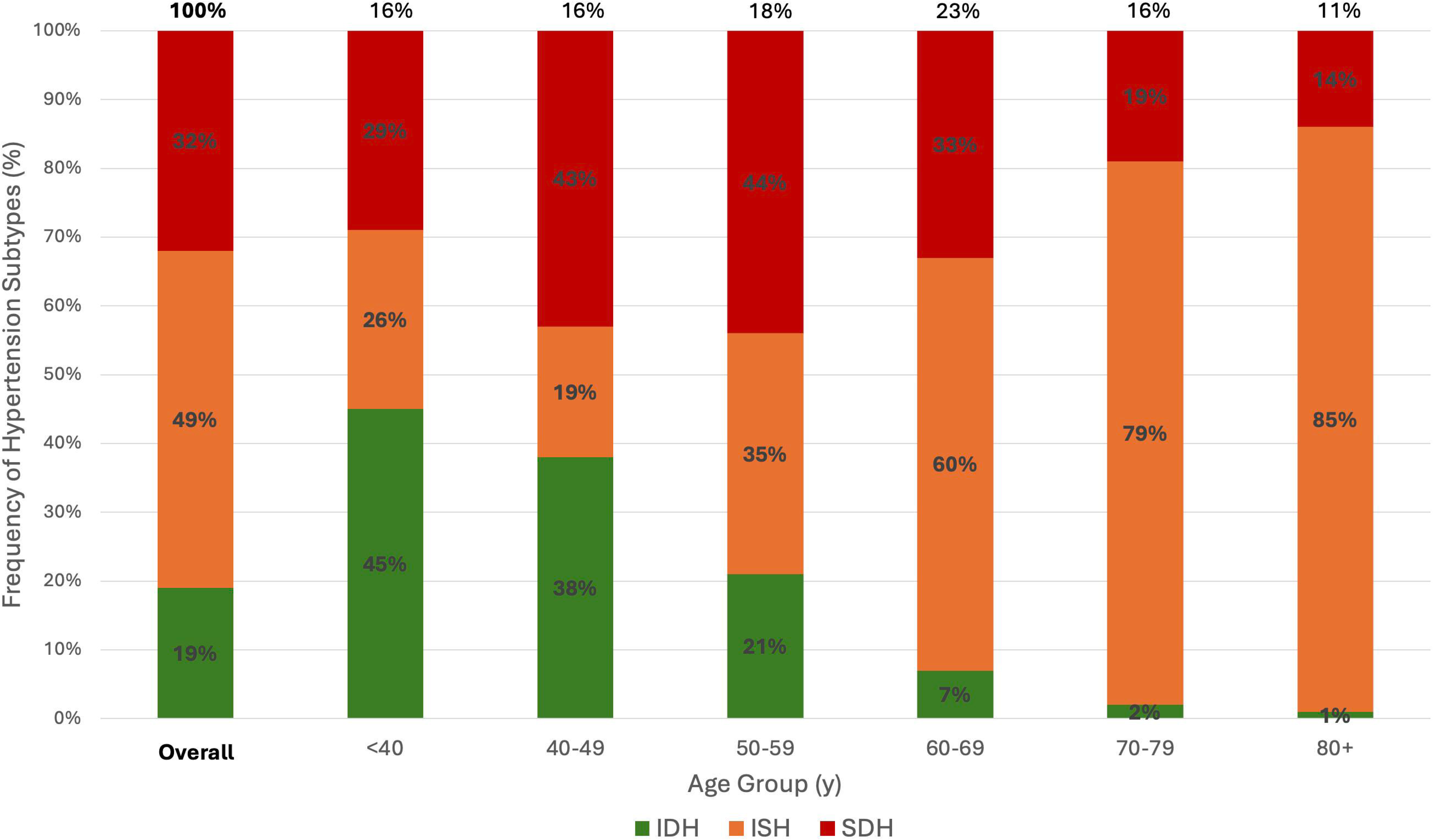
Prevalence of Hypertension Subtypes by Age in US Adults. ISH = Isolated Systolic Hypertension, IDH = Isolated Diastolic Hypertension, SDH = Systolic-Diastolic Hypertension

Untreated hypertension accounted for 30.5% of hypertensive individuals (projected to 13.8 million US adults). Another 45.5% of all hypertensive adults were treated to a blood pressure of < 130/80 mmHg (projected to 20.6 million US adults). Across increasing age, there was a decreasing prevalence of IDH, increasing prevalence of ISH, and increasing prevalence of SDH which peaked in the 6^th^ decade of life, but decreased afterwards. While IDH comprised over 40% of all HTN in those under age 40, ISH comprised over 80% of HTN in those aged 80 and over. Mild variation in HTN subtype prevalence among sexes was demonstrated in Figure 2; while the proportion of SDH was similar in all age groups, there was a trend towards more IDH in men and more ISH in women, particularly in the 6^th^ and 7^th^ decades of life. Figure 3 illustrated HTN subtype prevalence among age groups and ethnicities, showing that non-Hispanic Black and Hispanic demographics exhibited higher prevalence of ISH and SDH (and consequently lower prevalence of IDH) up until the 5^th^ decade of life, after which these differences attenuated significantly.

**Figure 2.**
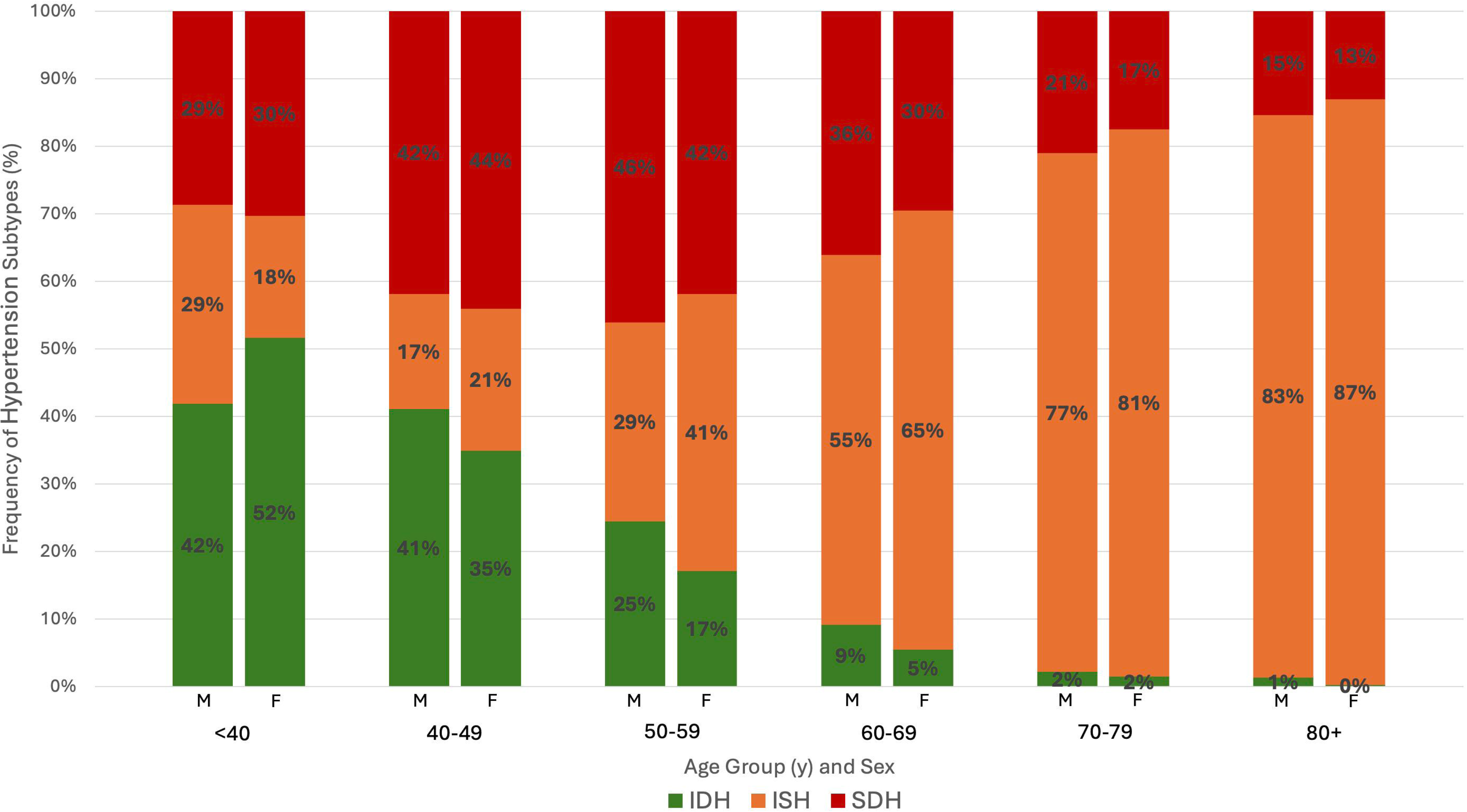
Prevalence of Hypertension Subtypes by Age and Sex in US Adults. M = Male, F = Female, ISH = Isolated Systolic Hypertension, IDH = Isolated Diastolic Hypertension, SDH = Systolic-Diastolic Hypertension

**Figure 3.**
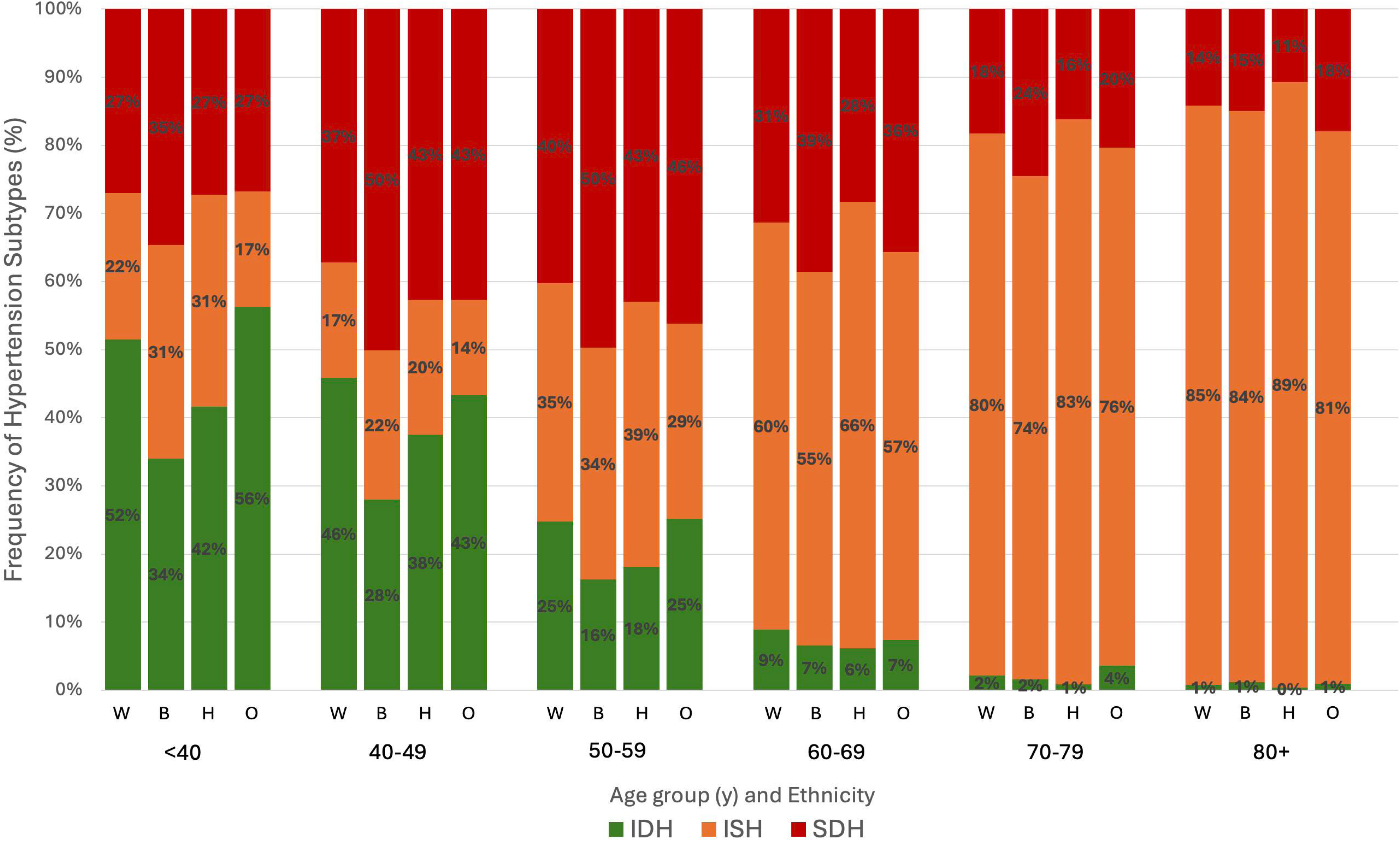
Prevalence of Hypertension Subtypes by Age and Ethnicity in US Adults. W = Non-Hispanic White, B = Non-Hispanic Black, H = Hispanic, O = Other ISH = Isolated Systolic Hypertension, IDH = Isolated Diastolic Hypertension, SDH = Systolic-Diastolic Hypertension

### Hypertension Subtypes as Predictors of Cardiovascular and All-Cause Mortality

For mortality analysis, the baseline cohort using NHANES data from years 1999-2008 included 23,206 subjects (projected to 194 million US adults). Of these, 58% were normotensive, 21% had ISH, 8% had IDH, and 13% had SDH. In adjusted mortality analyses compensating for age, sex, race/ethnicity, tobacco use, DM, dyslipidemia, and CKD, ISH was significantly associated with increased all-cause mortality (hazard ratio [HR]=1.13, 95% confidence interval [CI] 1.03-1.23, p<0.01) while SDH was associated with significantly increased CVD (HR=1.28, 95% CI 1.05-1.57, p=0.02) and all-cause mortality (HR=1.20, 95% CI 1.06-1.35, p<0.01) [Table 2]. IDH was associated with a protective effect for all-cause (HR=0.73, p<0.01) mortality in the unadjusted analysis, but when adjusted for the above factors there was no significant association with mortality risk. When adjusted further for use of antihypertensive therapy, all associations persisted with negligible change in HRs.

**Table 2.**
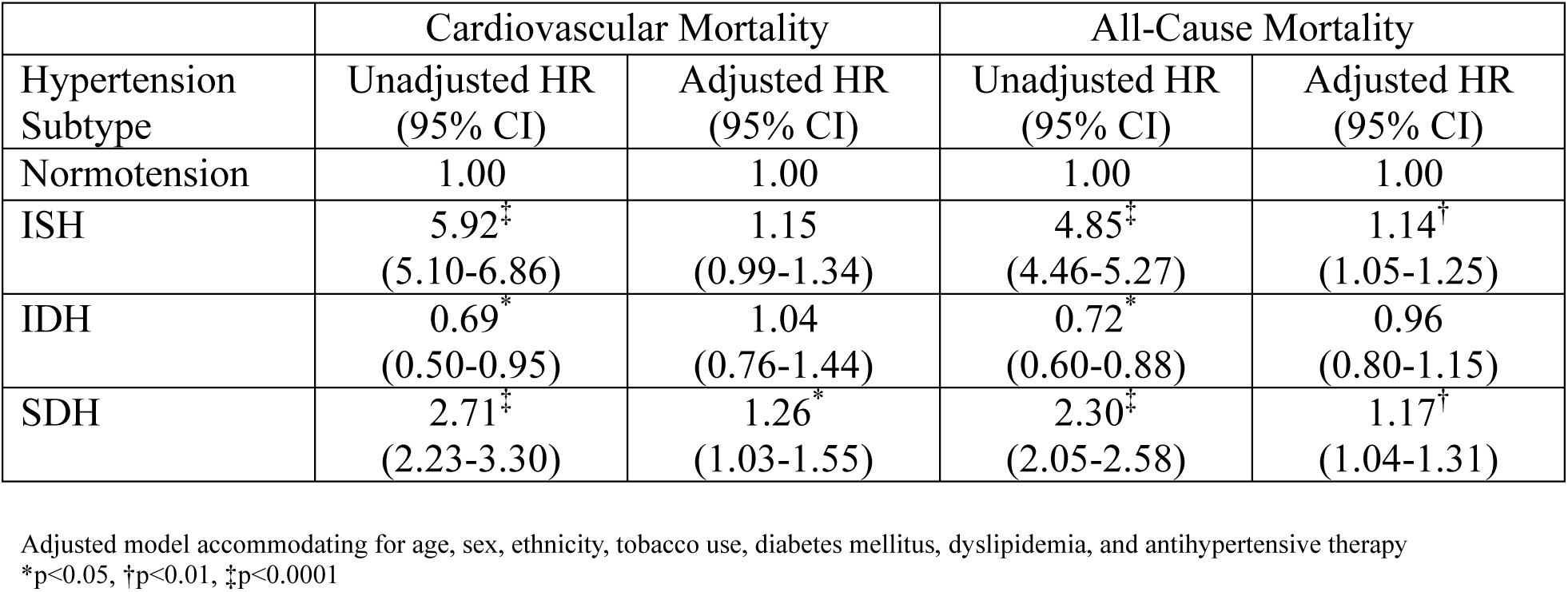
Unadjusted and Adjusted Hazard Ratios for Cardiovascular and All-Cause Mortality by Hypertension Subtype, NHANES 1999-2008.

When considering mortality risk in patients who were reclassified to HTN stages according to the 2017 ACC/AHA guidelines, elevated blood pressure (SBP 120-129 mmHg, DBP <80 mmHg) was not associated with increased CVD or all-cause mortality when adjusted for the above risk factors (p = 0.455 and 0.161, respectively) (Table 3). Individuals with stage I HTN did not have a significantly increased risk for CVD mortality (HR 1.13, p=0.161) but experienced significantly increased risk for all-cause mortality (HR 1.14, p=0.043) when compared to normotension.

**Table 3.** Unadjusted and Adjusted Hazard Ratios for Cardiovascular and All-Cause Mortality by Hypertension Stage, NHANES 1999-2008.

| Hypertension Stage | Cardiovascular Mortality |  | All-Cause Mortality |  |
| --- | --- | --- | --- | --- |
|  | Unadjusted HR (95% CI) | Adjusted HR (95% CI) | Unadjusted HR (95% CI) | Adjusted HR (95% CI) |
| Normotension | 1.00 | 1.00 | 1.00 | 1.00 |
| Elevated Blood Pressure | 1.89<sup>‡</sup><br>(1.52-2.34) | 1.08<br>(0.88-1.33) | 1.79<sup>‡</sup><br>(1.57-2.05) | 1.10<br>(0.96-1.25) |
| Stage I | 3.67<sup>‡</sup><br>(3.08-4.37) | 1.13<br>(0.95-1.35) | 3.22<sup>‡</sup><br>(2.85-3.63) | 1.14<sup>*</sup><br>(1.00-1.28) |
| Stage II | 7.90<sup>‡</sup><br>(6.41-9.74) | 1.40<sup>†</sup><br>(1.37-1.72) | 6.11<sup>‡</sup><br>(5.49-6.80) | 1.31<sup>†</sup><br>(1.17-1.47) |

Adjusted model accommodating for age, sex, ethnicity, tobacco use, diabetes mellitus, dyslipidemia, and antihypertensive therapy
\*p<0.05, †p<0.01, ‡p<0.0001

Adjusted Cox regression analyses by sex found no increased risk of CVD mortality in either the male (n=11,176, projected to 89 million) or female (n=12,003, projected to 95 million) cohorts for any HTN subtype compared to normotension. Women also did not demonstrate any increased all-cause mortality risk with any HTN subtype while men were found to have increased all-cause mortality risk with ISH (HR=1.17, 95% CI 1.02-1.35, p=0.03) and SDH (HR=1.26, 95% CI 1.07-1.48, p<0.01). In the adjusted subgroup analysis by ethnicity, ISH conferred increased risk for all-cause mortality only in Hispanic and non-Hispanic White individuals (HR=1.32, 95% CI 1.05-1.67, p=0.02; HR=1.13, 95% CI 1.02-1.25, p=0.02, respectively). Non-Hispanic Black individuals with SDH exhibited increased risk for both all-cause (HR=1.26, 95% CI 1.03-1.55, p=0.02) and CVD (HR=1.74, 95% CI 1.28-2.37, p<0.001) mortality, respectively.

## Discussion

The results of our analysis offer a detailed view of HTN subtype prevalence in the US adult population from 1999-2018 using contemporary definitions of HTN per the 2017 ACC/AHA HTN guidelines. We demonstrate that ISH remains the predominant HTN subtype in adults aged ≥50 years and is consistent with prior analyses including that of Franklin et al.^2^ from a much earlier cohort spanning NHANES years 1988 to 1994. We have further examined HTN subtype prevalences and mortality relationships according to sex and ethnicity. Redefinition of HTN treatment targets according to the 2017 ACC/AHA guidelines increases the total number of individuals with HTN (from a projected 42.7 million US adults (24%) per Franklin et al. to a projected 97 million US adults (41.1%) in our analysis), consistent with other contemporary analyses of the NHANES dataset.^12^ Compared to the earlier Franklin et al. analysis of NHANES III data^2^, our analysis using the 2017 ACC/AHA guidelines for HTN cutoffs decreases the prevalence of ISH (64.9% to 43%) and increases the prevalence of SDH (21.2% to 32%) and IDH (14% to 25%). Few studies report on the breakdown of HTN subtype prevalence in the US adult population, although our data demonstrates similar trends to other analyses from East Asian populations.^17^ Nearly half of all hypertensive adults in our analysis were successfully treated to a BP of <130/80 mmHg, demonstrating improving domestic awareness of HTN that is consistent with prior studies of HTN treatment and awareness trends both in the US^18^ and worldwide.^19,20^

To our knowledge, our subgroup analyses by sex and ethnicity also constitutes the first assessment of inter-group variance in mortality risk by HTN subtype. Men appear to have higher all-cause mortality risk with systolic HTN while women appear to be more protected from these effects. Previous work has already demonstrated significant differences in HTN prevalence, awareness, and BP control amongst ethnic groups.^21,22^ Our analysis further reveals a higher propensity for all-cause and CVD mortality in non-Hispanic Black US adults in particular with systolic HTN when compared to other ethnic groups. Persistent awareness and treatment disparities should be addressed in future research with special consideration of the role social determinants of health play in HTN prevalence and CVD outcomes.

SDH and ISH demonstrate significant association with cardiovascular and all-cause mortality while IDH is not significantly associated with either cardiovascular or all-cause mortality when adjusted for covariates. Systolic BP as the primary determinant of mortality risk in hypertensive individuals is well-documented for adults above the age of 50, although some data suggest that the mortality correlation for DBP is more pronounced in younger individuals.^23^ Other studies have demonstrated that widened pulse pressure characterized by DBP < 70 mmHg in the setting of ISH represents an independent risk factor for increased CVD events.^24–26^ In the clinical setting, special attention must therefore be paid to controlling both SBP and DBP with both lifestyle and pharmacologic interventions. In elderly patients with ISH, SBP reduction must be pursued with concomitant consideration for excessively low DBP or elevated pulse pressure, both of which may represent increased arterial stiffness and risk for CVD events. First-line antihypertensive agents should be initiated early and in combination with alternate agents for dual therapy if tolerated, with consideration for presence of comorbid conditions and certain drug combinations that have been demonstrated to be more effective in reducing CVD events compared to others.^27^ More research is needed to quantify the mortality risk for HTN subtypes in populations with specific comorbidities (eg, DM, CKD) that are independently associated with arterial stiffness and development of CVD.

Our observational study has several limitations. First, due to the nature of cross-sectional studies, associations demonstrated between covariates do not necessarily reflect causal relationships. Second, despite efforts to control for confounding variables through statistical methods, residual confounding remains a concern, as unmeasured or unknown factors (including antihypertensive therapy or other potentially relevant comorbidities that were not controlled for in our analysis) may influence the observed associations. However, it should be noted that most of the trends reported in this study are consistent with other analyses of both US and non-US populations. Finally, the subgroup analyses conducted for sex and ethnicity may be limited by subgroup sample size and thus not powered to detect smaller associations with all-cause and CVD mortality risk. Given the methods of the NHANES survey, we were also not able to control for other social factors including socioeconomic status and health literacy in our analysis.

Our analysis demonstrates using the revised 2017 ACC/AHA cut points for HTN that IDH remains the predominant HTN subtype in young persons, ISH the predominant subtype in older persons, with SDH peaking in the 6^th^ decade of life. ISH and SDH also remain most strongly predictive of CVD and total mortality. Further work is needed to demonstrate whether treatment approaches could be better informed by consideration of HTN subtype to improve CVD outcomes in those with HTN.

## Data Availability

NHANES data is publicly available online via the Centers for Disease Control and Prevention.

## Non-standard Abbreviations and Acronyms

ACC: American College of Cardiology
AHA: American Heart Association
BMI: Body Mass Index
BP: Blood Pressure
CI: Confidence Interval
CKD: Chronic Kidney Disease
CVD: Cardiovascular Disease
DALY: Disability-Adjusted Life-Year
DBP: Diastolic Blood Pressure
DM: Diabetes Mellitus
eGFR: Estimated Glomerular Filtration Rate
HR: Hazard Ratio
HTN: Hypertension
IDH: Isolated Diastolic Hypertension
ISH: Isolated Systolic Hypertension
NHANES: National Health and Nutrition Examination Survey
SBP: Systolic Blood Pressure
SDH: Systolic-Diastolic Hypertension

## Acknowledgements

No artificial intelligence tools or assistive writing technologies were used in the writing of this manuscript.

## Sources of Funding

Support was provided by the Stanley S. Franklin MD Memorial Endowment Fund for Hypertension and Heart Disease Prevention Research from the University of California, Irvine.

## Disclosures

Dr. Wong receives research funding not related to the current study from Novartis, Regeneron, and Novo Nordisk and is a consultant for Amgen, Novartis, Ionis and Heart Lung. The remaining authors disclose no conflicts of interest.

## Notes

### Competing Interest Statement

The authors have declared no competing interest.

### Author Declarations

IRB approval not required

